# COVID-19 outbreak response: a first assessment of mobility changes in Italy following national lockdown

**DOI:** 10.1101/2020.03.22.20039933

**Authors:** Emanuele Pepe, Paolo Bajardi, Laetitia Gauvin, Filippo Privitera, Brennan Lake, Ciro Cattuto, Michele Tizzoni

## Abstract

Italy is currently experiencing the largest COVID-19 outbreak in Europe so far, with more than 100,000 confirmed cases. Following the identification of the first infections, on February 21, 2020, national authorities have put in place an increasing number of restrictions aimed at containing the outbreak and delaying the epidemic peak. Since March 12, the whole country is under lockdown. Here we provide the first quantitative assessment of the impact of such measures on the mobility and the spatial proximity of Italians, through the analysis of a large-scale dataset on de-identified, geo-located smartphone users. With respect to pre-outbreak averages, we estimate a reduction of 50% of the total trips between Italian provinces, following the lockdown. In the same week, the average users’ radius of gyration has declined by about 50% and the average degree of the users’ proximity network has dropped by 47% at national level.

---

The COVID-19 outbreak was declared by the WHO a Public Health Emergency of International Concern on January 30, 2020 and since then, it has infected more than 650,000 individuals in more than 130 countries, reaching pandemic proportions (*1*). As of mid-March, Italy is by far the most affected country other than China that faced a sustained local transmission, which is putting a substantial burden on the Italian healthcare system and is threatening the socio-economic stability of the country.

Although there is a general consensus about the scarce efficacy of international travel restrictions to avoid case importation (*2, 3, 4, 5*), many governments have imposed partial or complete ban of air travel from Wuhan (*6, 7, 8, 9, 10*). Nevertheless, with an estimated 60% undetected cases (*11, 12, 13*) the novel coronavirus is currently circulating uncontrolled worldwide. Recent results suggest that a large fraction of infectious individuals are asymptomatic (*14*), dramatically shrinking the window of opportunity to contain local sporadic outbreaks by stopping chains of infection (*15*). Therefore, several European countries are now enacting non-pharmaceutical interventions aimed at slowing and mitigating the epidemic to “flatten the curve” and avoid the collapse of their entire healthcare system due to severe cases requiring hospitalization and extended stay in intensive care units (*16, 15, 17, 18*).

Italy has been the first EU country to implement unprecedented measures to restrict citizens’ mobility and try to contain the COVID-19 epidemic, rapidly escalating to more aggressive interventions to reduce social mixing and interrupt transmission chains. The policies introduced between the end of February and the first week of March ranged from school closure, advice against traveling or even banning non-authorized trips to and from areas with sustained transmission, university closure, ban of large-scale and public events, and then of any social gatherings, closure of museums, increasing restrictions on the opening hours of restaurants and bars, and encouraging or mandating smart/remote working whenever possible. Almost every day from February 25 to March 11, new and stricter policies have been declared in an increasing number of Italian provinces (see the Supplementary Material for a brief timeline). Since March 12, 2020, the whole country is under lockdown.

The mitigation measures enacted as part of the response to the unfolding COVID-19 pandemic are unprecedented in their breadth and societal burden. A major challenge in this situation is to quantitatively assess in real-time the impact of non-pharmaceutical interventions (NPIs) such as mobility restrictions and social distancing, to better understand the ensuing reduction of mobility flows, individual mobility changes, and their impact on contact patterns. A recent study has investigated the effect of NPIs deployed in China to contain the COVID-19 outbreak, finding that some of them, such as early case detection and contact reduction, appear to be the most effective (*19*).

To assess the impact of NPIs imposed by Italian authorities in response to the COVID-19 epidemic, we analyze de-identified, large-scale data from a location intelligence and measurement platform, Cuebiq Inc. This first-party data is collected from users who have opted-in to provide access to their location data anonymously, through a GDPR-compliant framework. Location is collected anonymously from opted-in users through a Software Development Kit (SDK) included in partner apps. At the device level, the operating system (iOS or Android) combines various location data sources (e.g., GPS, mobile network, WiFi networks, beacons) and provides geographical coordinates with an estimated accuracy level. Location accuracy depends on many factors and is variable in time, but it can be as accurate as 10 meters. Temporal sampling of anonymized users’ locations is also variable and depends on the operating system of the device and on user behavioral patterns, but it is generally high-frequency. We remove short-time dynamics by aggregating the data over 5-minute windows. The basic unit of information we process is an event of the form `(anonymous hashed user id, time, latitude, longitude)`, which we call a user’s *stop* in the remainder, plus additional non-personal metadata and location accuracy. We perform all our analyses on a panel of about 167,000 users who were active during the first week of the outbreak (February 22-28, 2020), i.e for whom there was at least one stop collected during that week. This led to about 175 million data points over the 8 weeks of this study.

We assess the effects of public health policies on the mobility and mixing patterns in Italy, by measuring (i) changes in the traffic flows of users between provinces; (ii) changes in the average distance traveled by users; (iii) changes in the spatial proximity of users. We average different mobility and proximity metrics during the pre-outbreak period (between January 18 and February 21, 2020) and observe their weekly and daily evolution as the intervention policies are enforced. To measure mobility networks, We create origin-destination (OD) matrices by measuring the number users who have traveled between any two Italian provinces over a given day, or a given week. A trip between two provinces *i* and *j* is recorded whenever a user makes two consecutive stops in *i* and *j* with more than 1 hour difference between the two. To characterize individual mobility patterns, we use the radius of gyration, *r*_*g*_, to quantify the mobility range of an individual during a given week (*20*). To measure the effects of social distancing, for every province we define a proximity network where nodes are individuals, and two individuals are connected by an edge if they made at least one stop within a distance *d* = 100 m from one other, within the same hour of the day. We then compute the average degree of such hourly proximity network, and average that over all the 1-hour slices of a given day, yielding a daily average degree ⟨*k*⟩ for every province. We use the daily time series of ⟨*k*⟩ to study how the proximity patterns defined above change as a consequence of mobility restrictions.

## Travel restrictions and mobility networks

The introduction of mobility restrictions across the country has significantly and progressively reduced the number of individuals moving between Italian provinces. After creating weekly OD matrices, we first look at the total number of users who moved out of a given province or entered from the rest of the country, in a week, defined as the total outgoing or incoming traffic, respectively. In the first week of the outbreak, from February 21 to 29, that we will call indifferently Week 1 from now on, following the establishment of “red zones” around areas with confirmed local transmission (Lodi, Cremona, Padova), all provinces in Lombardy, Piedmont and Veneto experienced a decrease in the outgoing number of users ranging between 21% and 43%. On the other hand, provinces in Central and Southern Italy exhibit a less marked reduction of mobility, consistent with the initial interventions focusing on the North of the country (Fig. 1A). During the same week we observe that for some provinces of the North, such as the Aosta Valley which is an important skiing resort area, the incoming volume of users slightly increased (+13%), suggesting that a non negligible fraction of users was still moving for recreational reasons at the time, or moving for self-quarantine outside of the major urban areas.

**Figure 1:**
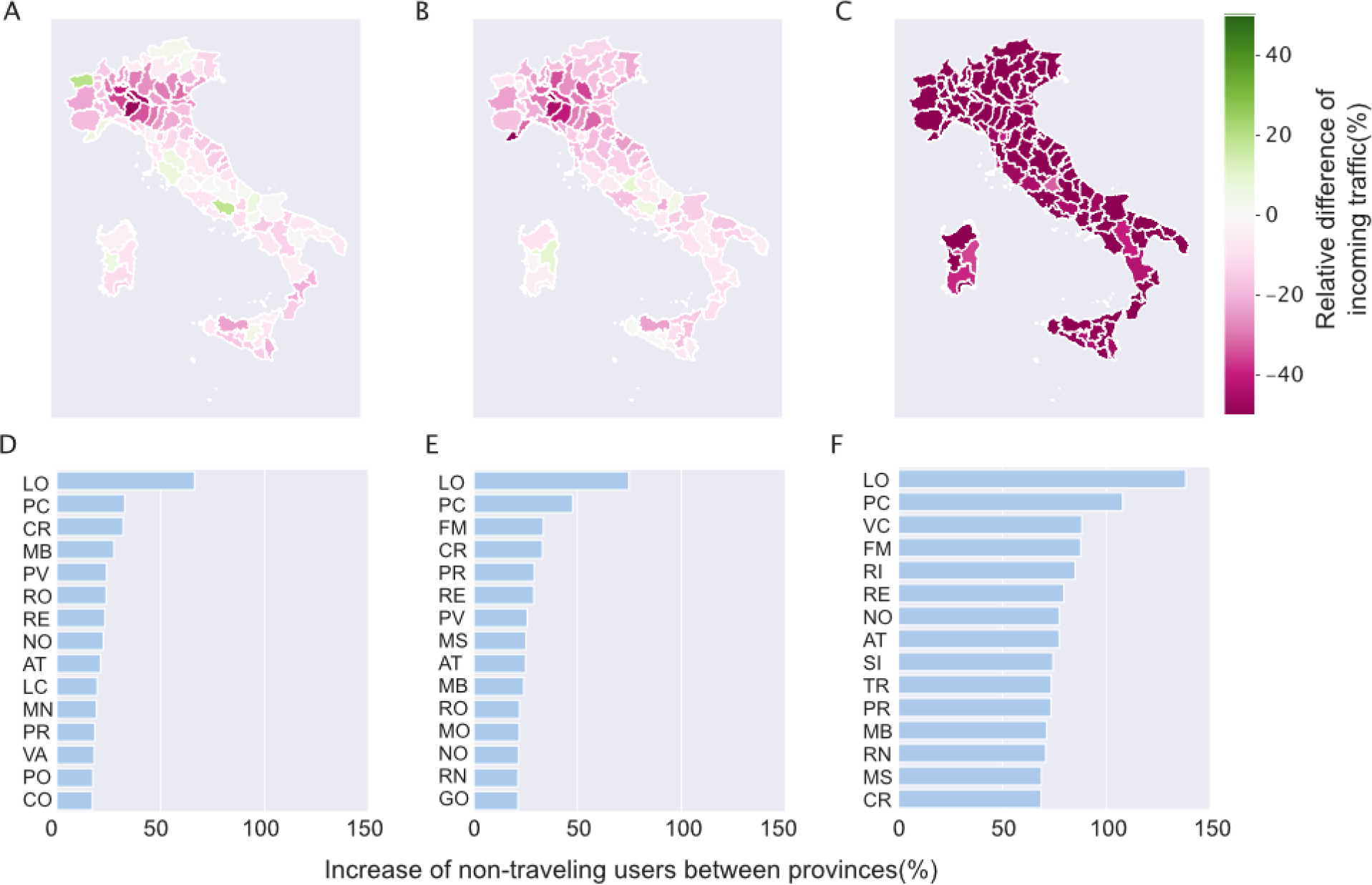
Effects of restrictions on Italian mobility. (**A**) Relative difference of the number of incoming users in each province during the first week of the outbreak (February 22-28), with respect to the number of incoming users average over the 5 weeks before the outbreak. (**B**) Same as (A) for the week of February 29 - March 6. (**C**) Same as (A) for the week of March 7-13. (**D**) Relative increase in the fraction of users who remain in a single province during the week of February 22-28. (**E**) Same as (D) for the week of February 29 - March 6. (**F**) Same as (D) for the week of March 7-13.

Between February 29 and March 6, that we will call indifferently Week 2 in the remainder, tighter restrictions on mobility across the provinces in Northern Italy have led to a reduction of the incoming traffic of about 17% across Piedmont, Lombardy, Emilia-Romagna and Veneto (Fig. 1B). Outgoing traffic decreased by 11% at the national level, maintaining marked regional differences with an average of 5% for the Southern provinces (see Fig. S1 in the Supplementary Information file). Finally, after March 7 (we will call Week 3 the week starting on that day), the enforcement of a national lockdown has led to a deep and almost uniform reduction of the outgoing and incoming traffic between all provinces by an average 40% at national level (Fig. 1C).

The fraction of users who remained in a unique province increased by an average of 9%, at the national level, during the first week of the outbreak, by 13% in the second week and by 54% in the week of the lockdown. Initially, the proportion of stationary users has increased more than 50% only in the province of Lodi, where the first COVID-19 outbreak was detected (Fig. 1D,E). On Week 3, such figure markedly increased across the whole country, and in some provinces it more than doubled with respect to the pre-outbreak period (+138% in Lodi and +108% in Piacenza), as reported in Fig. 1F.

By further inspecting the changes in the number of people traveling along specific connections, we first look at the connections that had more than 200 weekly travelers before the outbreak. On those links, we observe an average reduction in the number of travelers of 16% in Week 1, 13% in Week 2, and 41% in Week 3. On a number of connections, especially between provinces in Northern Italy, the reduction has been particularly high since the Week 1 (Fig 2A). For instance, among the links with the largest decrease in travelers, we note the presence of the connection Turin-Milan, facilitated by a fast train line usually taken by daily commuters. The observed 50 to 80% reduction in the users traveling on that line, from Week 1 to Week 3, is likely a consequence of the remote-working policy adopted by many companies. Similar arguments apply to the reduction observed for many other connections such as Milan-Novara (−56%), Genoa-Alessandria (−55%) and Milan-Como (−54%).

**Figure 2:**
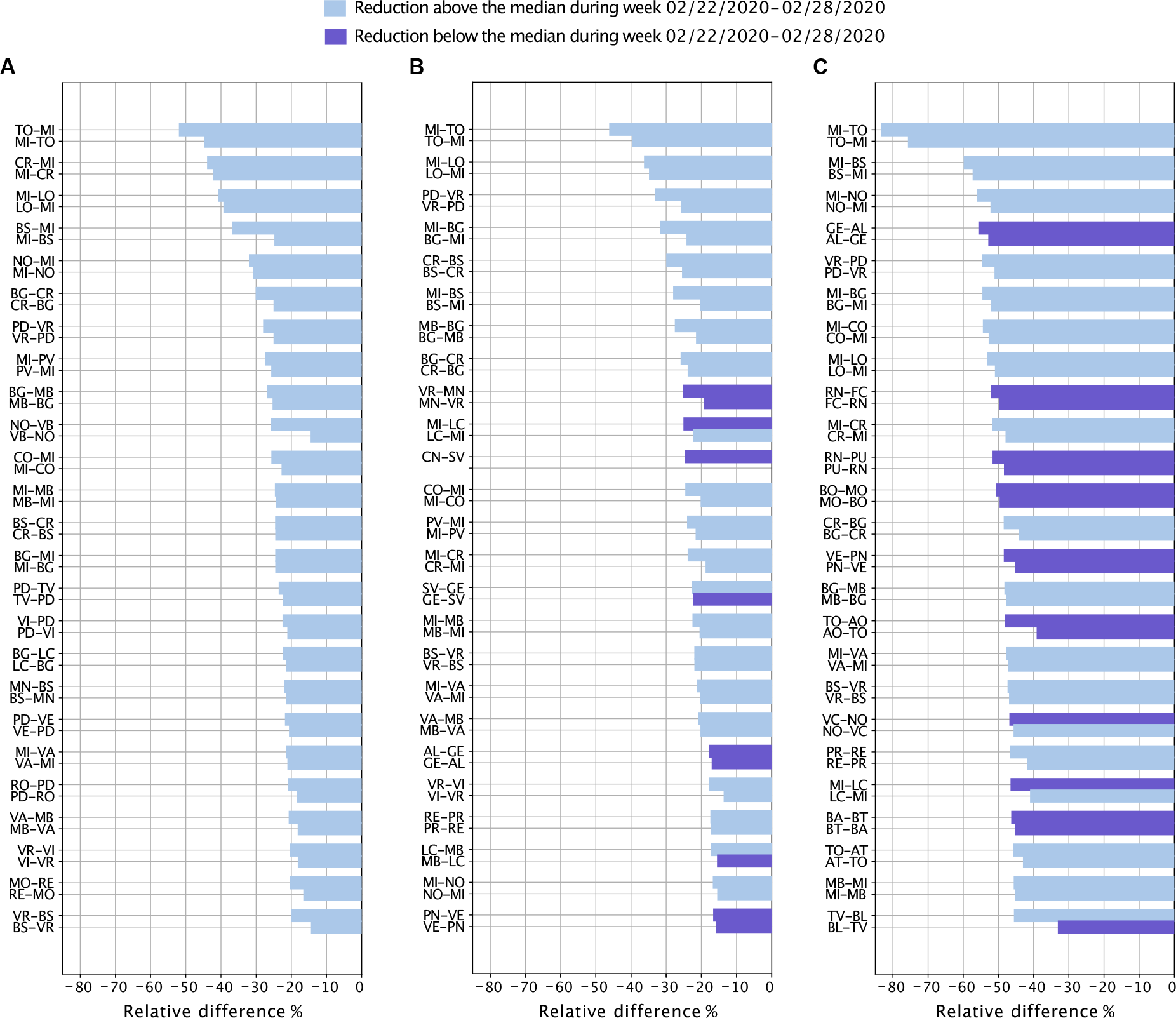
Effects of the restrictions on mobility network connections. Top 50 connections between Italian provinces by relative difference in the number of travelers observed during Week 1 (**A**), Week 2 (**B**) and Week 3 (**C**). We only show connections with an average number of traveling users *≥* 200 in the pre-outbreak period. Darker color indicates the connections whose reduction in traveling users was below the median in Week 1.

Thanks to the high granularity of the data under study, we are also able to inspect changes in the short-range mobility behavior of the users. Specifically, we measure the individual weekly radius of gyration, *r*_*g*_, for each user and we compute distributions of its values over all users who are resident in a given province. The home province of a user is defined as the province where the majority of a user’s stops were recorded at night (12 AM - 6 AM) as in Ref. (*21*) over the 5 weeks before February 21, 2020. By analysing variations in the weekly radius of gyration, we find a significant reduction in the characteristic distance traveled by individuals, starting on February 22, right after the initial COVID-19 outbreak was identified in Lodi. In Fig. 3, we report the median weekly radius of gyration, 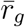, in the Italian provinces, comparing the value of 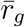 measured before the outbreak and during the outbreak. Even though mobility restrictions were enforced only in Lombardy and neighbouring regions, we observe that 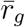 has decreased in all provinces of Italy since the very beginning of the outbreak. Initially, as expected, we observe the largest percent decrease in the provinces of Lodi (LO), Imola (IM), Cremona (CR), at the center of the outbreak (Fig. 3A). However, significant reductions are observed in some Southern provinces, like Taranto (TA) or Messina (ME), as well. As more restrictions were put in place, the average radius of gyration by province has further declined across the country, with an average 49% reduction in Week 3, from 13 km to 7 km, at national level. In Week 3, half of the users’ population typically travelled less than 2 km, while before the outbreak half of the population was traveling more than 5.7 km a week (see Fig. S2 in the Supplementary Information file). In particular, we observe the largest relative reduction of 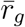 (from 6.1 km to 1.3 km) in the province of Milan (MI) which did not appear among the top 20 provinces in the precedent weeks (Fig. 3C).

**Figure 3:**
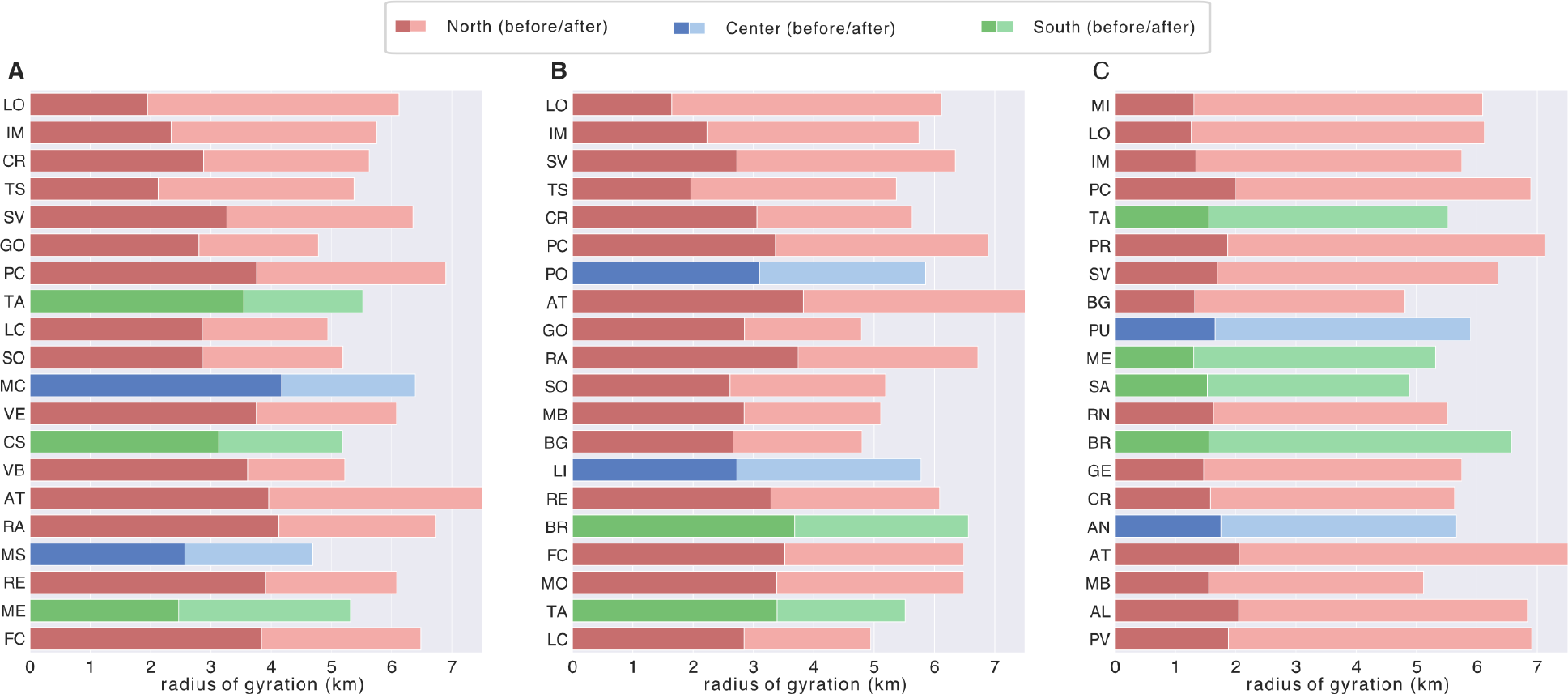
Individual mobility patterns: reduction of spatial range. Median radius of gyration for different Italian provinces, computed before and after the outbreak for Week 1 (**A**), Week 2 (**B**) and Week 3 (**C**). Provinces are ordered, top to bottom, by relative variation of the median radius of gyration from the pre-outbreak period (maximum relative variation at the top). Only the 20 provinces with the largest relative *r*_*g*_ variation are shown for each week.

## The effect of social distancing on proximity patterns

The average contact rate, or the number of unique contacts made by a person on a typical day is a fundamental quantity to model and understand infectious disease dynamics (*22*). Measuring social contacts that are relevant for disease transmission trough geo-location data, like the ones used in this study, is challenging, however, it is possible to measure proximity between users in the spatial range accessible by GPS accuracy (*23*). We consider changes in the spatial proximity of our users’ sample as an indicator that can be used to quantify the level of social distancing adopted by the population.

We evaluated the effect of NPIs on the proximity patterns of our users’ sample, by defining a proxy of the potential encounters each user could have in one hour. To this aim, we built a proximity network among users based on the locations they visited and the hour of the day when these visits occurred. To create such a network, we assert proximity between any two users in the same province who were seen within a circle of radius *R* = 50 m in a 1-hour period. The procedure to build the proximity network is described in Fig. 4A. If two users have been in proximity several times in a day, this still counts as only one link in the network. To measure variations in the network structure over time, we look at the average degree of the network defined as ⟨*k*⟩ = 2*E/N*, where *E* is the number of links and *N* is the number of nodes of the network. It is important to remark that this is not a close-range contact network. Rather, it captures a looser notion of closeness at the chosen spatial and temporal scales. A link between two nodes indicates the possibility that the corresponding individuals have had a close-range encounter during a given day. A sensitivity analysis on the spatial radius *R* and the temporal binning shows that the estimated reduction of the average degree is robust to variations of such parameters (see Table S1 in the Supplementary Information file).

**Figure 4:**
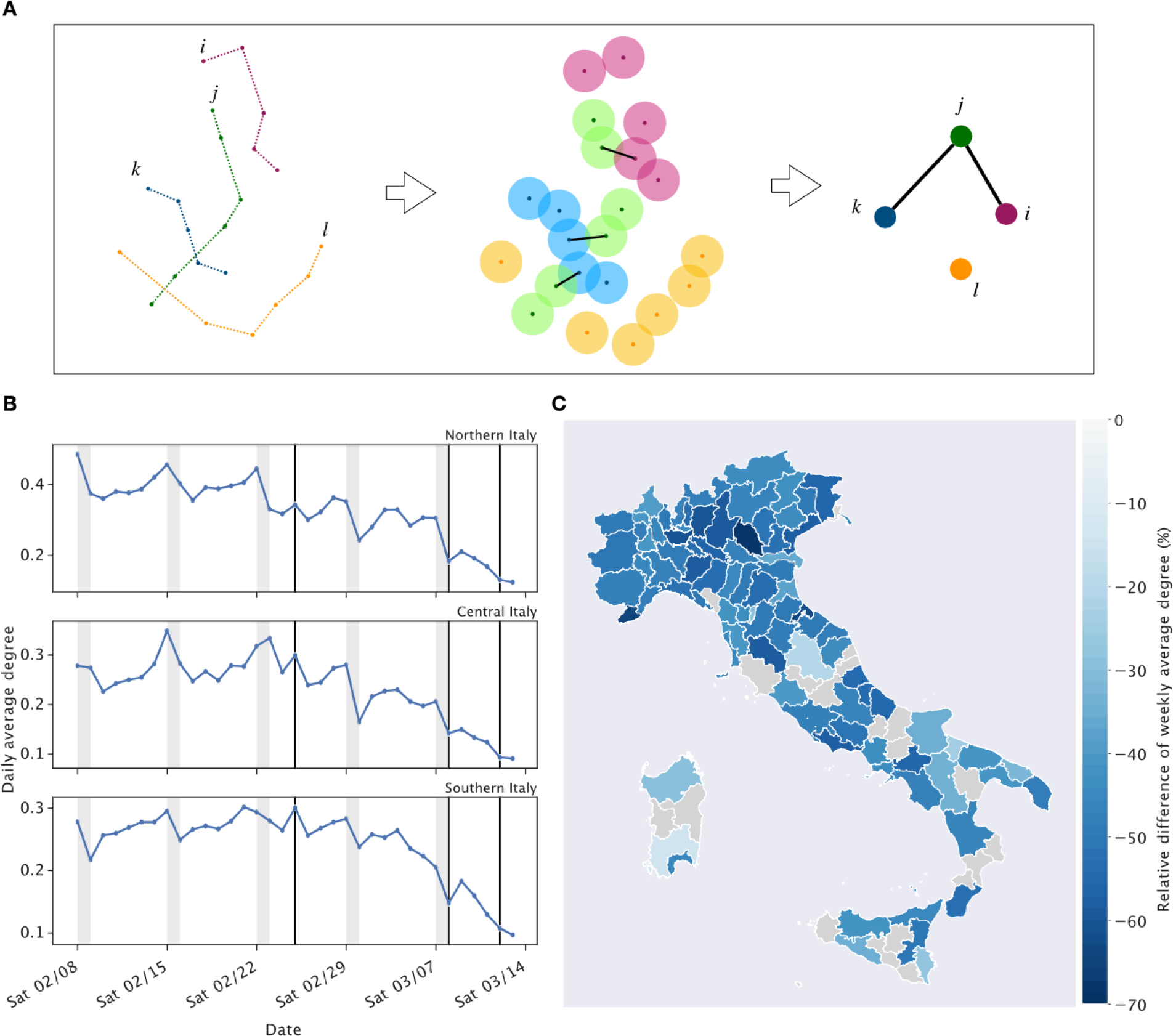
The effect of NPIs on the network of potential encounters. (**A**) For every users’ stop within a 1-hour time interval, a circle with radius 50m is drawn. If the circles belonging to two distinct users intersect, a link representing a potential interaction between them is built. (**B**) Time series of the daily average degree of the potential encounter network aggregated by the three Italian macro-regions. Solid black lines mark the major intervention policies implemented during the outbreak. Shaded grey areas highlight weekends. (**C**) Relative difference in the weekly average degree measured during Week 3 with respect to the pre-outbreak average, by province. The gray provinces correspond to areas where the sample is smaller than 500 users.

Fig. 4B shows how the daily average degree has changed over time in the Northern, Central and Southern Italy. In the Northern and Central Italy we observe a declining trend in ⟨*k*⟩ starting immediately after the first interventions took place on February 24. In the Southern Italy, we still observe a declining trend but with the presence of some peaks or a milder reduction in ⟨*k*⟩ during the first week of outbreak. This may be explained by the fact that southern regions were far from the center of the outbreak and there were still few or no reported cases in the area, at that time. When looking at all provinces of Italy, a consistent picture emerge: during the first week of outbreak the effects of social distancing on the users’ network were visible only in the Northern provinces and much less elsewhere (see Fig. 5 and Fig. S3 in the Supplementary Information).

**Figure 5:**
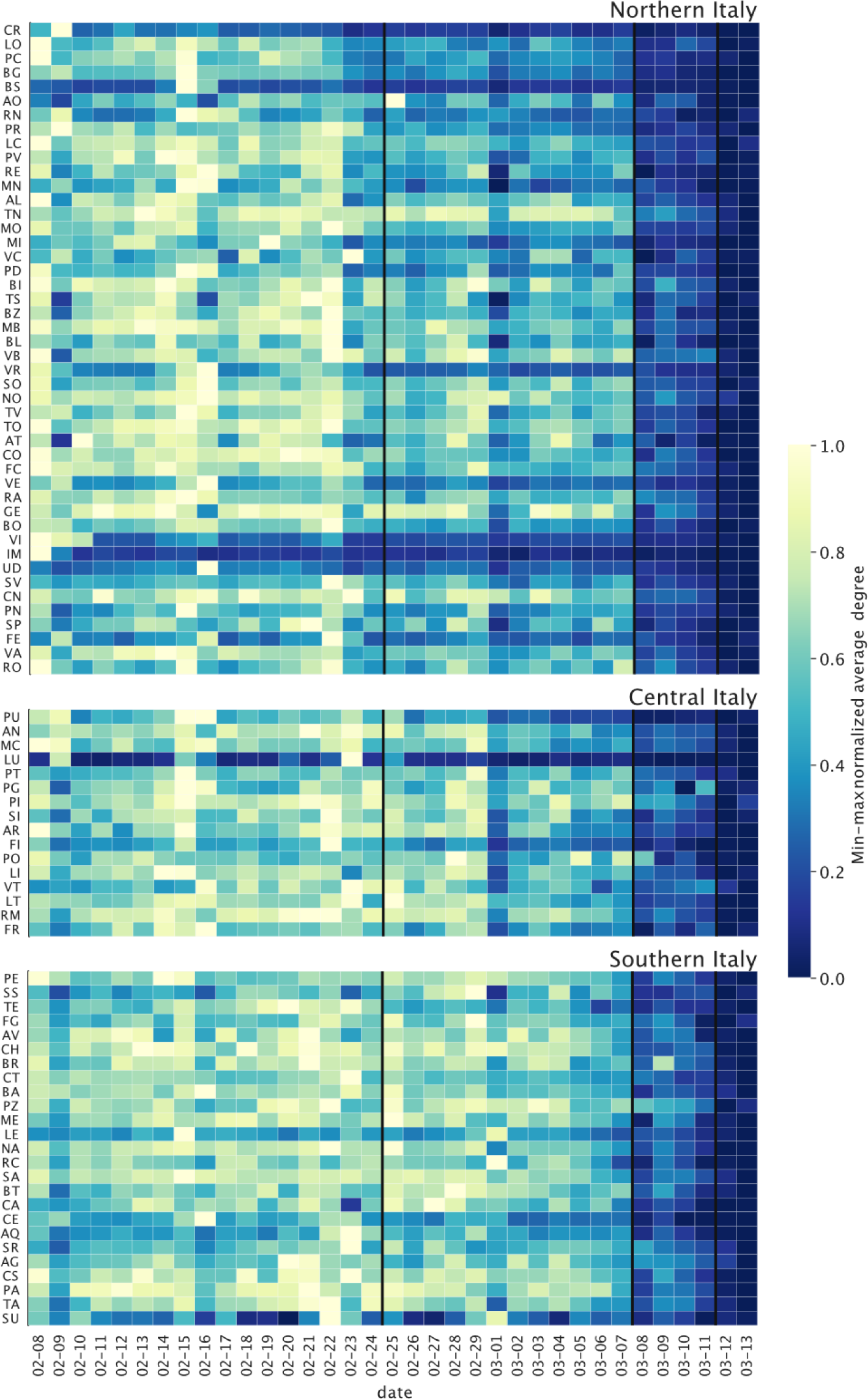
Spatio-temporal patterns of networks of potential encounters across Italian provinces. Time series of the average degree of the potential encounter network normalized between minimum and maximum value by province. Solid black lines mark the major intervention policies implemented during the outbreak.

We observe a consistent reduction of ⟨*k*⟩ across the whole country, only after February 29, 2020, when tighter movement restrictions were imposed in Northern Italy. Although still localized, such interventions were followed by an average 13% reduction of ⟨*k*⟩ across provinces, with respect to the pre-outbreak period. In Week 3, the average degree of the users’ proximity network further declined by 47%, but with significant differences within the country. Fig. 4C displays the relative difference in ⟨*k*⟩ between the third week of outbreak and the pre-outbreak average, for all provinces. In some provinces, such as Milan and Verona, ⟨*k*⟩ has declined by more than 60% when compared to the pre-outbreak averages. In some other parts of Italy, such as the regions of Sardinia and Umbria, we observe a much lower reduction, in the range from 10 to 25%. Such discrepancy might be due to differences in the number of reported COVID-19 cases by province, which may cause a different perceived risk of infection and consequently a limited adoption of protective behavior. On the other hand, it is important to notice that in some provinces we can measure the behavior only of a relatively small number of users, which limits our conclusions. In Fig. 4C, we don’t report values of ⟨*k*⟩ for provinces that have less than 500 users in our panel.

Finally, by assigning a geohash home location to each user (see Materials and Methods), we can also measure the relative fraction of proximity events that are observed within the home geohash and those that took place away from home. In Fig. S4 we report the fraction of proximity events observed away from a user home geohash, averaged over all users at national level. We clearly see a significant drop in such quantity - from 65% to 45% - right after March 8, 2020, when the government issued a nationwide stay-at-home guidance, thus highlighting the immediate impact of such policy on the Italian population.

## Discussion

In the coming months several countries will need to implement their epidemic response plans facing the unprecedented challenges posed by COVID-19. This will put an exceptional burden on society as a whole, with unexplored trade-offs between individuals’ freedom and coordinated actions for public interest.

Italy has been the first Western country where national authorities have escalated containment measures up to enforcing a nation-wide lockdown, and thus it represents an exceptional setting to assess the impact of such measures before other countries possibly take the same path. Here we show that during the early phase of the outbreak when only mild - yet socially expensive - measures were in place (e.g., school closure) the traffic towards/from the most affected provinces declined by about 50%. It is worth noticing that a mild drop in mobility was observed in the entire country, with a slight increase only in a few provinces, suggesting that individuals started to change their mobility behavior even beyond the official guidance, probably aiming to reduce the risk of getting infected. We observed similar results by also considering different mobility metrics: both the median radius of gyration and the average degree of the users’ proximity network consistently decreased all over Italy, with a few exceptions. Finally, when stricter measures were in place at the national level, the population reacted uniformly across the country, largely limiting trips between different provinces.

Our study comes with limitations. Although the sample of users under study well matches the population distribution of the Italian provinces (see Fig. S5 in the Supplementary Information file), it is not representative of the general Italian population in terms of age and socio-economic status although we verified that it does represent fairly well population distribution at the provincial level. The population under study is not constant over time because users can opt-out sharing their geo-location at any time. Also, the internal logic of the data gathering system might fail in sensing users who stop moving for a prolonged period of time. This suggests that our estimates may underestimate the true effects of mobility restrictions. Finally, we do not try to characterize the trips made by our users, and we have no information as to whether a given trip is among those with an authorized purposed. Therefore, our results should not be interpreted as an assessment of the adherence by the Italian population to the restrictions.

Because of the exceptional nature of the unfolding events, there is little available evidence to compare our results to. Recent studies, based on the analysis of digital traces and contact surveys, have demonstrated that NPIs were effective in containing the COVID-19 outbreak in China (*8, 19, 24*). The estimated maximumum reduction in mobility and social contacts in China during the lockdown was about 80%, with respect to a baseline set on January 1, 2020 (*19*). Such level of social distancing was sufficient to achieve control of COVID-19. We can not tell yet for how long the observed reductions in mobility and social mixing in Italy will be sustained, and whether they will prove sufficient to contain the COVID-19 outbreak. However, our findings indicate that real-time monitoring of mobility changes following NPIs is feasible through the analysis of de-identified geo-positioning data and it might be used to guide intervention policy over the course of the outbreak.

## Data Availability

The raw geo-located data analysed in the study are not publicly available due to privacy reasons.
All the data underlying the figures are deposited on the Humanitarian Data Exchange at the following URL: https://data.humdata.org/dataset/covid-19-mobility-italy.

## Author contributions

PB, LG, CC, and MT developed the idea and research. FP and BL collected the mobility data. EP, PB, LG, FP, and MT processed the mobility data. PB, LG, and MT wrote the first draft of the manuscript and all other authors discussed results and edited the manuscript.

## Data and materials availability

The raw geo-located data analysed in the study are not publicly available due to privacy reasons. All the data underlying the figures are deposited in the Humanitarian Data Exchange COVID-19 Outbreak page: https://data.humdata.org/dataset/covid-19-mobilityitaly.

## Supplementary Materials

Materials and Methods

Figures S1-S5

Tables S1-S2

References 1-6.

